# Somatic mutations in Parkinson disease are enriched in synaptic and neuronal processes

**DOI:** 10.1101/2020.09.14.20190538

**Authors:** Irene Lobon, Manuel Solís-Moruno, David Juan, Ashraf Muhaisen, Federico Abascal, Paula Esteller-Cucala, Raquel García-Pérez, Maria Josep Martí, Eduardo Tolosa, Jesús Ávila, Raheleh Rahbari, Ferran Casals, Tomas Marques-Bonet, Eduardo Soriano

## Abstract

The role of somatic mutations in complex diseases, including neurodevelopmental and neurodegenerative disorders, is becoming increasingly clear. To explore their relevance in sporadic Parkinson disease, we performed whole-exome sequencing in blood and four brain regions of ten patients. We identified 59 candidate somatic single nucleotide variants (sSNVs) through sensitive calling and extensive filtering. We validated 27 of them with amplicon-based deep sequencing, with a 70% validation rate for the highest-confidence variants. Most of the sSNVs were exclusively called in blood but were also found in the brain tissues with the ultra-deep amplicon sequencing, demonstrating the strength of multi-tissue sampling designs. We could confirm between 0 and 6 sSNVs per patient and generally those with a shorter lifespan carried more variants. Remarkably, the validated sSNVs are enriched in genes with synaptic functions that are co-expressed with genes previously associated with Parkinson disease.

## Introduction

Somatic mutations appear during development and tissue maintenance, making every individual a mosaic of cells with slightly different genomes. Early mutations occurring before gastrulation are shared by tissues of different germ layer origin (Bae et al. 2018; Lodato et al. 2015) and can cause disease (Mensa-Vilaró et al. 2019; Kluge et al. 2019). Mutations occurring later in development or during adult tissue maintenance that confer a proliferative advantage produce clonal expansion of the cells carrying them, limited by each tissue’s dynamics (Lee-Six et al. 2019; Moore et al. 2020). Notably, this type of somatic mutations are the cause of cancer (Nowell 1976), but have also be found in healthy skin (Martincorena et al. 2015; Abyzov et al. 2017), esophagus (Martincorena et al. 2018), colon (Lee-Six et al. 2019), liver (Brunner et al. 2019), endometrium (Moore et al. 2020) or lung (Yoshida et al. 2020). Further, organ-exclusive somatic mutations in the MTOR pathway are involved not only in cancer (Guertin and Sabatini 2007) but also in neurodevelopmental disorders such as hemimegaloencephaly (Poduri et al. 2012) and focal cortical dysplasia (Lim et al. 2015). Other studies linking somatic mutations to disease did not determine whether the variants were of early or late origin, as in a case of congenital arrhythmia where the causal mutation was only tested in cardiomyocytes and lymphocytes and found in both cell types (Priest et al. 2016).

Before birth, each human neuron carries 200 to 400 somatic single nucleotide variants (sSNVs) (Bae et al. 2018) and about 300 to 900 sSNVs can be detected by early infancy (Lodato et al. 2018). Those with a frequency higher than ~2% can also be detected in tissues that originate from a different germ layer, suggesting that brain development does not heavily rely on clonal expansion. Other types of somatic variation have also been found in neurons: retrotransposon mobilization is common and disproportionately impacts protein-coding loci (Baillie et al. 2011) and over three copy number variants (CNVs), mainly losses, can be found in each cell (Cai et al. 2014). Strikingly, while the number of CNVs decreases in brain with age (Chronister et al. 2019), sSNV load increases (Hoang et al. 2016; Lodato et al. 2018) posing questions about their relevance in neuronal diversification, plasticity and dysfunction.

The somatic variant hypothesis for neurodegenerative diseases states that unexplained sporadic cases could be caused by somatic mutations, presumably in the same genes affected in familial cases (Pamphlett 2004). Supporting this theory, age-related sSNVs accumulate faster in neurodegeneration (Lodato et al. 2018). However, not only these later-acquired mutations seem relevant, but also earlier somatic mutations may contribute to phenotypes and diseases. For example, individuals with Autism spectrum disorder have a higher burden of somatic mutations than their unaffected siblings, measured in blood (Dou et al. 2017). In Alzheimer, targeted sequencing of blood samples showed that somatic variants in autosomal dominant genes (such as *APP)* can explain ~2% of cases (Nicolas et al. 2018). Another study on blood and hippocampus exomes from 52 Alzheimer patients showed that over a fourth carried somatic mutations affecting pathways known to contribute to tau hyperphosphorylation (Park et al. 2019), demonstrating the power of exome analysis.

The link between Parkinson disease (PD) and somatic mutations is not as clear. Notably, only about 10% of cases can be attributed to monogenic forms (Lesage and Brice 2009) and at most 30% of patients have an affected first-degree relative (Rocca et al. 2004). A study on 511 sporadic cases tested multiple brain regions with a sensitivity limit at 5% of variant allele frequency (VAF) and did not find any somatic variants in *SNCA*, the main causative gene in early onset familial PD (Proukakis et al. 2014). On the other hand, patients showed high levels of heteroplasmic mitochondrial DNA deletions (Bender et al. 2006) and more *SNCA* somatic copy number gains in substantia nigra neurons compared to controls, which positively correlated with age of onset (Mokretar et al. 2018).

To explore the potential link between somatic SNVs and Parkinson disease, we sequenced the exome of five different tissues from ten sporadic Parkinson patients – substantia nigra, striatum, neocortex, cerebellum and blood – at an average coverage of 60X. We developed and implemented a filtering approach based on both single tissue and joint information which we used to identify 59 candidate sSNVs. Further amplicon-based deep sequencing confirmed 27 of them, with an average of 3 sSNVS per individual (range 0-6). The confirmed sSNVs were enriched in synaptic and axonal processes and patients with more sSNVs tended to die earlier, suggesting a potential role of these variants in the disease. Interestingly, over 76% of the variants validated in multiple brain tissues were only called in blood in the exome data, demonstrating that studying other more accessible and unaffected tissues may well serve for the identification of variants with lower frequency in diseased organs.

## Results

### Dataset

We sequenced the whole exome of five different tissues from ten sporadic Parkinson patients at an average coverage of 60X. Blood was obtained from stored vials while the cerebellum, neocortex, substantia nigra and striatum samples were collected during autopsies (Figure 1A). Individuals’ median age was 81 at the time of death, they had varying ages at disease onset and both sexes were represented (Supplementary table 1). As expected, germline SNV calling showed a good clustering of tissue samples by individual except for two samples from individual DV2, which appear to be contaminated (Supplementary figure 1). Accordingly, this sample was excluded from subsequent analyses but used for noise profiling.

**Figure 1.**
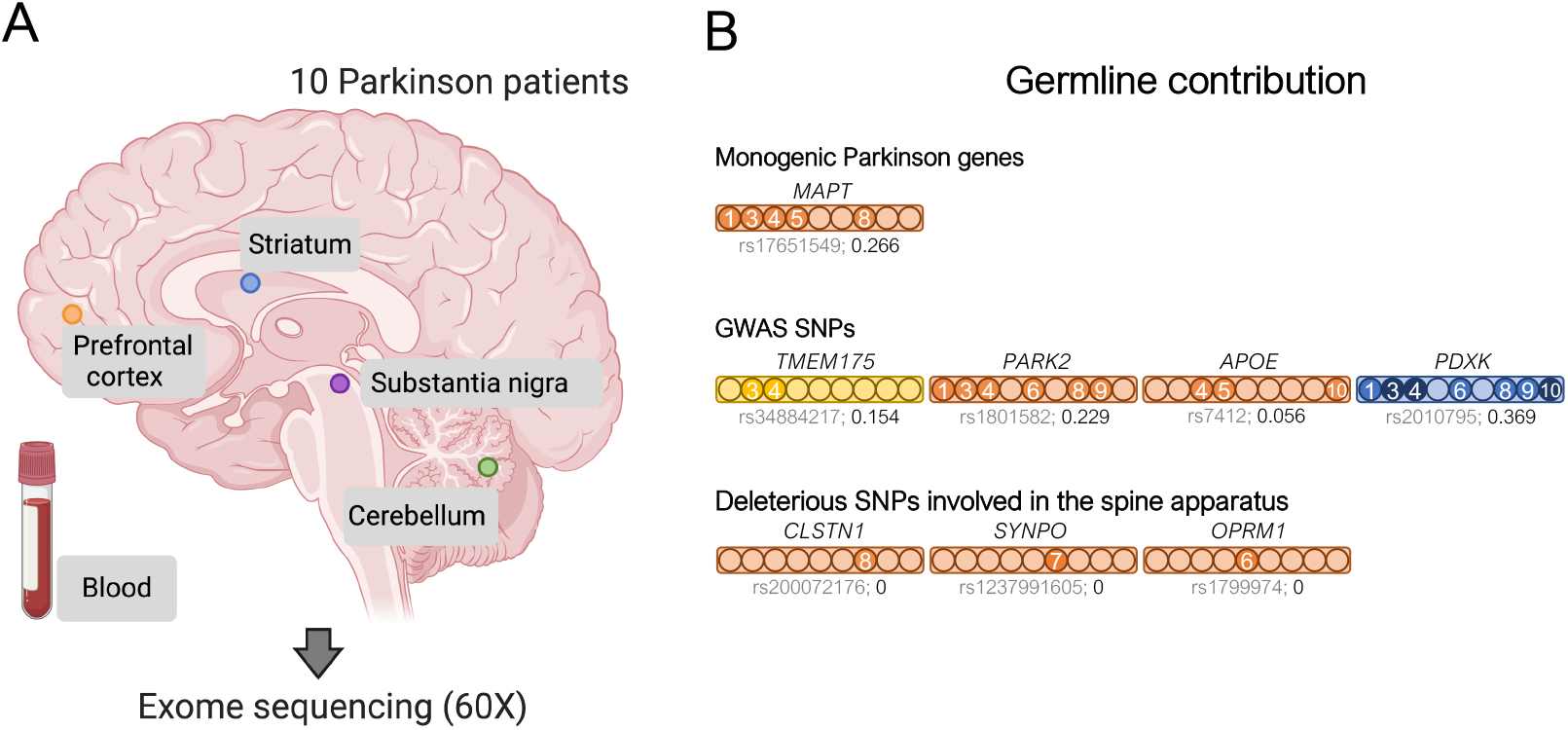
**A. Experimental design**. DNA was obtained from blood and four brain regions from ten sporadic Parkinson disease patients. Each sample’s exome was captured and seguenced. **B. Germline SNVs related to Parkinson**. A bar is shown for each germline variant and carrier individuals are indicated by filled circles (DV2 is not included). Darker circles indicate a homozygous SNV. The affected gene is indicated on top and the dbSNP identifier and the freguency of the alternative allele in the IBS population are shown below each bar. Bar color indicates the type of variant: missense (red), splice acceptor (yellow) or intronic (blue).

### Germline variants are associated with the disease

The subject individuals were categorized as sporadic Parkinson cases because no affected first-degree relatives were known. This assessment did not include genetic analyses, so we evaluated their germline variants before considering their somatic mutational landscape. We used three different strategies to prioritize germline single nucleotide changes. First, we identified germline SNVs in genes linked to Parkinson in OMIM (www.omim.org) that were deleterious as indicated by a CADD score > 15 (Rentzsch et al. 2019). Only one variant met these criteria, rs17651549, a missense mutation in the *MAPT* gene, which encodes the tau protein, predicted as deleterious by multiple methods and at a highly conserved position in vertebrates (Supplementary table 2). This variant has been previously linked to PD by different means: multivariate family-based association tests (Wang, Mullersman, and Liu 2010), pathway analysis (Song and Lee 2013) and targeted resequencing (Spataro et al. 2015). However, in a contradictory haplotype association analysis it provided a reduced risk for PD (J. Li et al. 2018). This SNP is not infrequent in Europe; particularly, the frequency of the alternative allele in the 1000GP IBS population (Iberian populations in Spain) is 0.27 and similarly, it was 0.28 in our Spanish cohort (Figure 1B).

Then, we made use of GWAS studies, with greater statistical power, to recognize Parkinson-associated germline variants (41 variants, Supplementary table 3) (Tan et al. 2010; Do et al. 2011; International Parkinson Disease Genomics Consortium 2011; International Parkinson Disease Genomics Consortium and Wellcome Trust Case Control Consortium 2 2011; Lill et al. 2012; Nalls et al. 2014a; Chen et al. 2016) and found four in our cohort. DV3 and DV4 carried a splice acceptor variant at *TMEM175*, rs34884217. This variant, predicted to affect nonsense-mediated decay, has been associated with Parkinson (Nalls et al. 2014; Heckman et al. 2017) and *TMEM175* deficiency is linked to the increase of α-synuclein aggregation (Jinn et al. 2017), indicating a possible causal link. The missense mutation of *PARK2* showed the highest odds ratio for Parkinson disease in Europeans in a meta-analysis (Ramakrishnan et al. 2016). The other two SNVs have been associated with the disease (Huang, Chen, and Poole 2004; Williams-Gray et al. 2009; Elstner et al. 2009), but conflicting results have also been reported (Federoff et al. 2012; Guella et al. 2010).

In addition, all germline SNVs were filtered by deleteriousness (CADD > 15 and SIFT prediction) and frequency in the 1000GP European population (< 0.1). The resulting 214 variants affected 207 genes, with each patient carrying a median of 27 variants (range 20-32). An overrepresentation enrichment analysis (Zhang, Kirov, and Snoddy 2005) found 7 molecular function and cellular component terms significantly enriched in these genes (FDR ≤ 0.05, Supplementary figure 2). Remarkably, the term with the highest enrichment ratio was *spine apparatus*, a derivate of the smooth endoplasmic reticulum generally present in dendritic spines that seems to participate in spine remodeling in Parkinson disease models (Smith, Villalba, and Raju 2009). *Kinesing binding* and *motor activity were* also among the significantly enriched terms, with 7 out of the 9 individuals carrying a deleterious mutation in genes driving these 3 associations. These genes have been previously linked to PD in different studies. As an example, *CLSTN1* overlapped a significantly hypomethylated CpG (Chuang et al. 2017), was differentially expressed (Kong et al. 2018) and carried a missense mutation (Yemni et al. 2019) in PD cases. Several dynein and kinesin proteins also appear to be relevant in PD. All of this highlights the complexity of Parkinson disease, where common and rare variants affect multiple pathways that seem to contribute to the phenotype. Abundant data is needed to uncover these associations, so the evaluation of somatic mutations has the potential to contribute to this effort.

**Figure 2.**
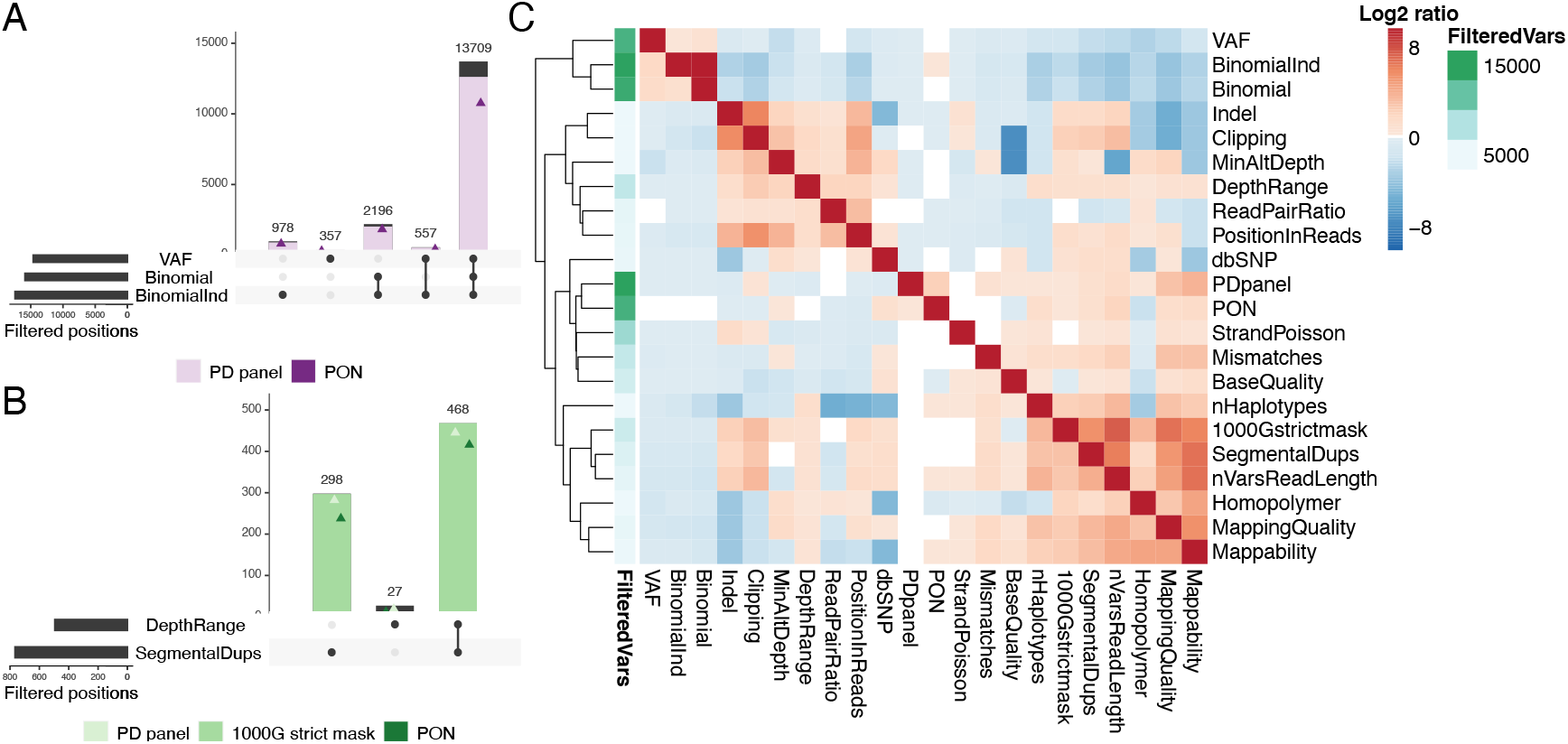
**Relevance of COSMOS filters in our Parkinson exome data**. **A. Germline SNVs**. Intersection of the main filters that identify germline heterozygous point mutations: VAF (high variant allele frequency), Binomial (non-significant binomial test for allele depths) and BinomialInd (non-significant binomial test in at least one sample from the individual). For each intersection, the number of variants also found in the other individuals of our dataset (PD panel, filled in lavender) and an external panel (PON, purple triangle) are shown. **B. CNV filters**. Intersection of DepthRange (most extreme depth variants) and SegmentalDups (segmental duplications WGAC track) with PD panel and PON (light and dark green triangles, respectively) and variants present in the 1000GP strict mask filled in green. **C. Relationship between filters**. Log2 ratio between the number of variants failing both the row and the column filter and those failing the row filter only. Red indicates higher log2 ratios and hence higher co-appearance of filters. The annotation column indicates the total number of positions failing each of the row filters.

### Somatic variant calling

Somatic variant calling was first developed for cancer and standard approaches are based on paired tumor and normal samples comparison (Krøigård et al. 2016; Xu 2018). Growing evidence shows that low frequency somatic mutations are present in multiple tissues, even from different germ layers (Bae et al. 2018; Lodato et al. 2015), so paired comparisons are inadequate in scenarios with no or limited clonal expansion. Requiring two supporting reads in our 60X data would imply a mean VAF of over 3%. These variants could have originated before gastrulation, be present in multiple tissues and require joint sample calling. On the other hand, blood clonal expansion, especially prevalent in old age (Jaiswal et al. 2014), will result in variants private to this tissue, requiring single sample calling. In standard paired samples calling, shared calls are essentially discarded, which filters out germline variants and recurrent artefacts. The main obstacle unpaired single or joint sample calling have is precisely differentiating these confounding factors from true somatic variants.

Germline callers are optimized to discard variants that do not fit with set VAF expectations. Low frequency variants, such as those we are interested in, are considered sequencing noise or contamination and discarded. For this reason, we called variants in all our samples using HaplotypeCaller with —ploidy 10, which increases its sensitivity to lower frequency variants. We also used VarScan 2 with lax parameters (--min-coverage 1 --min-reads2 1 --p-value 1 --min-var-freq 0.000001). The resulting call set is mostly composed of germline heterozygous SNVs, calls within CNVs, recurrent sequencing errors and other artefacts. Hence, we developed a filtering strategy to identify high confidence somatic SNVs, COSMOS (Combined Or Single sample MOSaicism detection, https://github.com/ilobon/COSMOS). COSMOS can be used to annotate the relevant information needed to filter somatic candidates on standard VCFs as well as to filter calls directly. Filtering can be performed with a single sample approach, a joint filtering approach using multiple samples from the same individual, or both.

### COSMOS

Manual inspection of the calls obtained from HaplotypeCaller and VarScan 2 allowed the identification of multiple sources of artefacts, for which we devised filtering approaches. The main rationale was that even if our patients had the same disease, and could carry mutations in the same pathways or even genes, the probability they would bear mutations in the exact same position was negligible in just 10 individuals. Hence, variants present in multiple individuals are presumably artefacts.

We discarded all off-target calls because they have higher strand imbalances (69.4% of off-target calls fail the Poisson test vs. 39.5% of on-target calls) and read pair imbalances (21.8% vs 4.4% failing the read pair ratio filter). The most frequent confounding factors for on-target calls were germline heterozygous SNVs, CNV regions, unresolved regions of the genome and regions that are more difficult to align (indels, homopolymers).

Germline heterozygous calls supposed at least 93.1% of our on-target calls. They can be easily distinguished when their VAF is close to or higher than 50%. However, germline calls’ VAFs are more over dispersed the lower the coverage (Supplementary figure 3) and VAFs as low as 18% can result from germline variants sequenced at 60X (and even 10% for 20X, our lower bound depth). We used a binomial test (83.2% of on-target calls) and a VAF upper limit (76.5% of on-target calls had VAF > 40%) to identify the most obvious cases, with a large overlap between the two sets of calls (Figure 2A). These variants are common to all tissues from the same individual, so when multiple samples are available, requiring that all pass the binomial test greatly increases the detection power, following an exponential distribution (Supplementary figure 4). We found an additional 5.5% of variants with this approach (first bar in Figure 2A). However, an alternative way of flagging most of these calls is by a large number of called individuals. By using the other 45 samples from the remaining 9 PD individuals, which results in a batch and population-specific panel, we can identify 92.1% of them. Alternatively, an external panel of normals (PON) consisting of 428 whole genomes sequenced at the Sanger Institute tagged 82.6% of them (Figure 2A).

Regions with CNVs can create artefactual somatic variants when reads are collapsed -mapped into a single copy of the reference genome. CNV callers can be used to identify the highest confidence regions, but more sensitive approaches are needed to obtain high-confidence somatic variant calls. We excluded regions with extremely low or high coverage (keeping 75% of the distribution) or overlapping the WGAC segmental duplications track, which filtered out 60.4% of the remaining on-target non-germline calls. 97.5% of these calls overlap the 1000GP strict mask, which identifies regions of the genome with recurrently higher/lower coverage or lower mapping quality, showing its suitability as an alternative. Again, most calls failing our CNV filters were called in multiple individuals (94.1% in our PD panel and 84% in the independent PON) (Figure 2B). Other features useful to identify these artefacts are a high number of variants within read length distance, an imbalance in the number of mismatches in reads carrying each allele or in the proportion of clipped reads and a low mappability (Supplementary figure 5).**C**

Unresolved regions of the genome or highly variable duplications can result in collapsed mapping with no significant increment of coverage because only reads spanning the homologous region and carrying few variants will be mapped. These are easily identified by a biased position of the alternative allele in the reads, clustered at read ends. We defined a position-in-reads bias score (PIR) to address this. Other features targeting these artefacts are allele clipping imbalances, a high number of variants in the vicinity, more than three haplotypes, allele imbalances for mapping quality and read pair imbalances (Figure 2C). Local alignment around indels and homopolymers is challenging and technical and biological noise are difficult to distinguish, so we discarded close-by variants to increase our call set confidence (Methods). Finally, variants with too few reads supporting the alternative allele cannot be distinguished from random noise and their imbalances cannot be evaluated, so a hard cutoff was applied (minimum of two and three reads supporting the alternative allele for joint and single sample calling respectively).

### Most somatic validated variants are present in both blood and brain

A total of 59 variants passed COSMOS filtering, 7 of which were called exclusively by VarScan. After manual inspection, we classified variants in four tiers of confidence (examples in supplementary figures 6 and 7). Tier 1 had 17 high confidence variants, tier 2 consisted of the 7 VarScan calls, tier 3 contained 29 lower confidence candidate sSNVs and tier 4 included 5 putative false positives (Supplementary table 4 and Methods). A median of 7 candidates were called in each individual (range 1-11) and perhaps unsurprisingly given the limited coverage, 91.5% of all the candidate variants were called in just one tissue, mostly blood (61.1% of single tissue calls) (Figure 3A).

**Figure 3.**
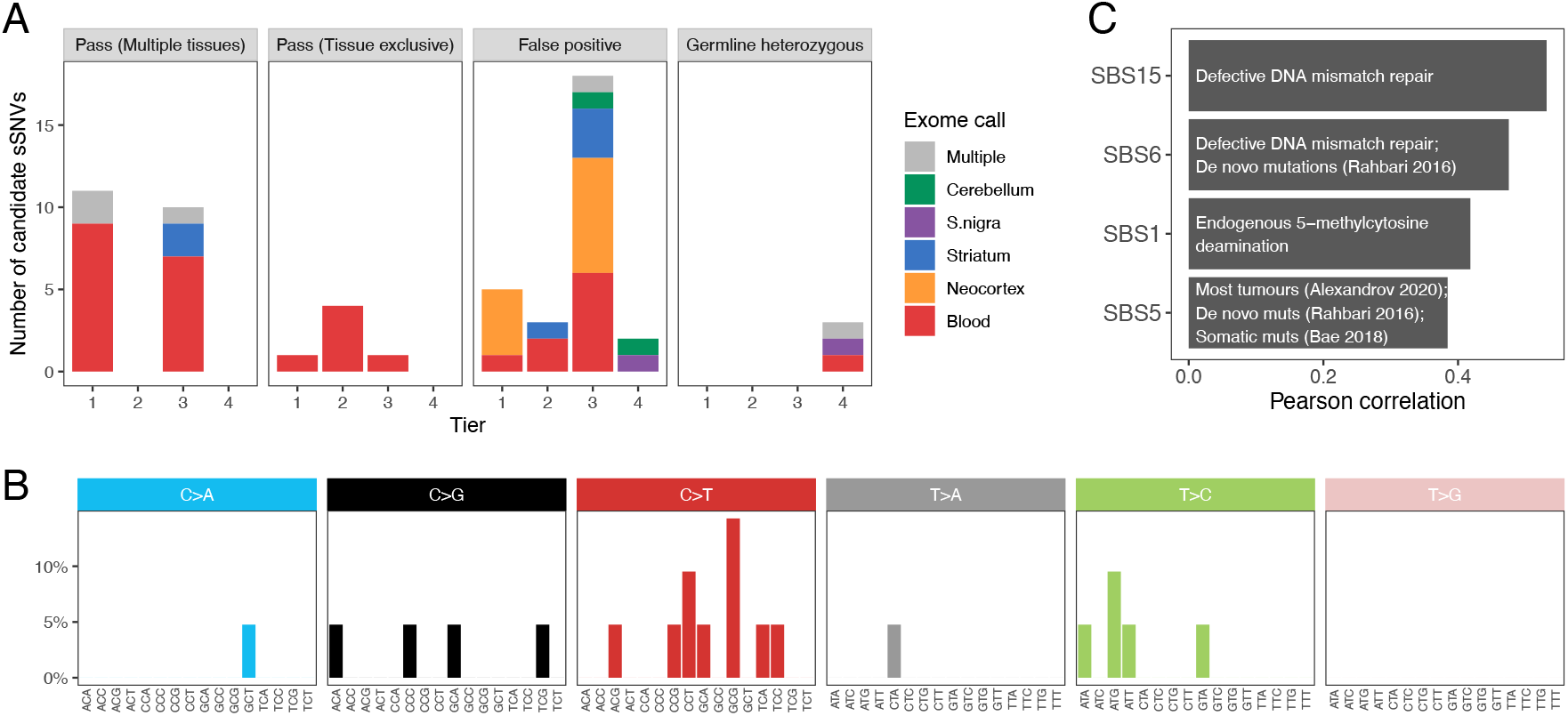
**A. Validation of candidate variants**. Number of mutations validated (Pass) in multiple or a single tissue, or found to be false positives or germline heterozygous variants with amplicon sequencing data. Variants are distributed by calling-confidence tier and colors indicate the tissue in which the variant was originally called in the exome data. **B. Mutational spectrum of somatic SNVs in Parkinson brains**. Variants present in the brain of Parkinson patients, segregated by substitution and trinucleotide context. **C. Correlation with COSMIC signatures**. Moderate Pearson correlations (r>0.3) between the spectrum of brain somatic variants and the single base signatures (SBS) from COSMIC. Text describes the etiology or studies relevant for each signature.

We evaluated 58 out of the 59 variants with the rhAmpSeq targeted amplicon sequencing system, which allows the multiplexed amplification of multiple genomic positions. This way, we sequenced data from all positions and all patients at a mean coverage of over 18,000X so that non-called individuals could be used as negative controls. Four of the samples could not be amplified enough and had low coverages (mean ≤ 2,300 X, Supplementary figure 8). To consider a variant as a validated sSNV in a given sample, we required that (1) it was the second most common allele in that sample and (2) its VAF was higher than the mean + 2 standard deviations of all other individuals’ samples VAFs with sufficient coverage (defined as not lower than the mean – 1 standard deviation). We additionally classified variants passing these filters in multiple tissues of an individual and with VAFs > 30% in all of them as germline heterozygous variants (Supplementary material).

A total of 27 somatic SNVs were validated using these criteria. Validation rates were 70.6% for tier 1 (12 out of 17), 57.1% for tier 2 (4 out of 7), 37.9% for tier 3 (11 out of 29) and 0% for tier 4 (0 out of the 5 negative control variants). Per individual, a median of 4 variants were validated and the median validation rate was 50% (mean of 40.6%, ranging from 0% to 83%) as all calls from 3 individuals (DV4, DV5 and DV9) were false positives. DV9 had just one candidate sSNV call but the other two had 7 and 6, respectively, demonstrating moderate interindividual variability. Only 6 variants were present in a single tissue, which was always blood (Figure 3A, second panel). Interestingly, 76.2% (16 out of 21) of variants validated in multiple tissues had been called exclusively in blood in the exome data (Figure 3A, first panel), as they have a higher frequency in this tissue (mean difference of 10.8%, mean VAF in blood 11.5% and 0.7% in the other tissues). This was not a consequence of our calling method, as 68.8% of these variants (11 out of 16) had no read supporting the alternative allele in any brain sample but were then validated in at least 2 brain regions. This probably results from their random amplification in tissue maintenance of blood but could also be a consequence of depletion in the central nervous system.

### The brain somatic SNVs spectrum is most similar to the expected mutational signatures

The discovery that different mutagenic agents –such as UV light, carcinogens or intrinsic cell processes– produce distinct substitution patterns in a context dependent manner led to the development of mutational signature analysis (Alexandrov et al. 2013). To obtain the mutational spectrum of Parkinson disease, we combined the 21 sSNVs validated in at least one brain sample and classified them by substitution and trinucleotide context (Figure 3B). As the number of variants was insufficient for mutational signature deconvolution, we calculated its Pearson correlation with the COSMIC single base signatures. Since most of the identified variants are present in multiple tissues and are therefore of early origin, we expected to find a high similarity to signatures SBS1 and SBS5. Indeed, we found both among the four moderate correlations (Pearson’s r > 0.3, Figure 3C). SBS1 is an ubiquitous signature that results from the spontaneous deamination of methylated cytosines. SBS5 can be detected not only in most cancer samples (Alexandrov et al. 2020) but also in *de novo* mutations (Rahbari et al. 2016), somatic mutations (Bae et al. 2018) and in population level variants (Mathieson and Reich 2017). Interestingly, SBS6 was also found to be highly correlated with the *de novo* mutational spectrum (Rahbari et al. 2016). The other signatures we found to be similar to the Parkinson spectrum, SBS15, has been proposed to be caused by defective DNA repair.

### Variant allele frequency of sSNV can be used to cluster the tissues

We validated sSNVs in 6 out of the 9 subject individuals, ranging from 1 to 6 variants per patient, a modest yet sensible number for an exome analysis. All of DV10’s sSNVs were only detected in blood but the other 21 variants (from DV1, DV3, DV6, DV7 and DV8) were detected in at least two brain tissues. We clustered the tissues based on their VAFs at these sSNVs. Because each individual has just a few variants, we pooled them together to analyze general tissue dynamics (Figure 4A). As expected from its clonal expansion, blood is the most distant tissue, with brain tissues being more closely clustered. Remarkably, striatum and substantia nigra, both affected in Parkinson disease (Bernheimer et al. 1973), cluster together, which could be caused by their closer developmental origin and/or more similar physiology. The presence of 3 variants with higher frequencies in all brain tissues than blood suggests that our findings are not the result of blood contamination in the other tissues. Importantly, blood was not called in the exome sequencing data for these variants, and the amplicon sequencing confirmed the VAF distribution in the different tissues.

**Figure 4.**
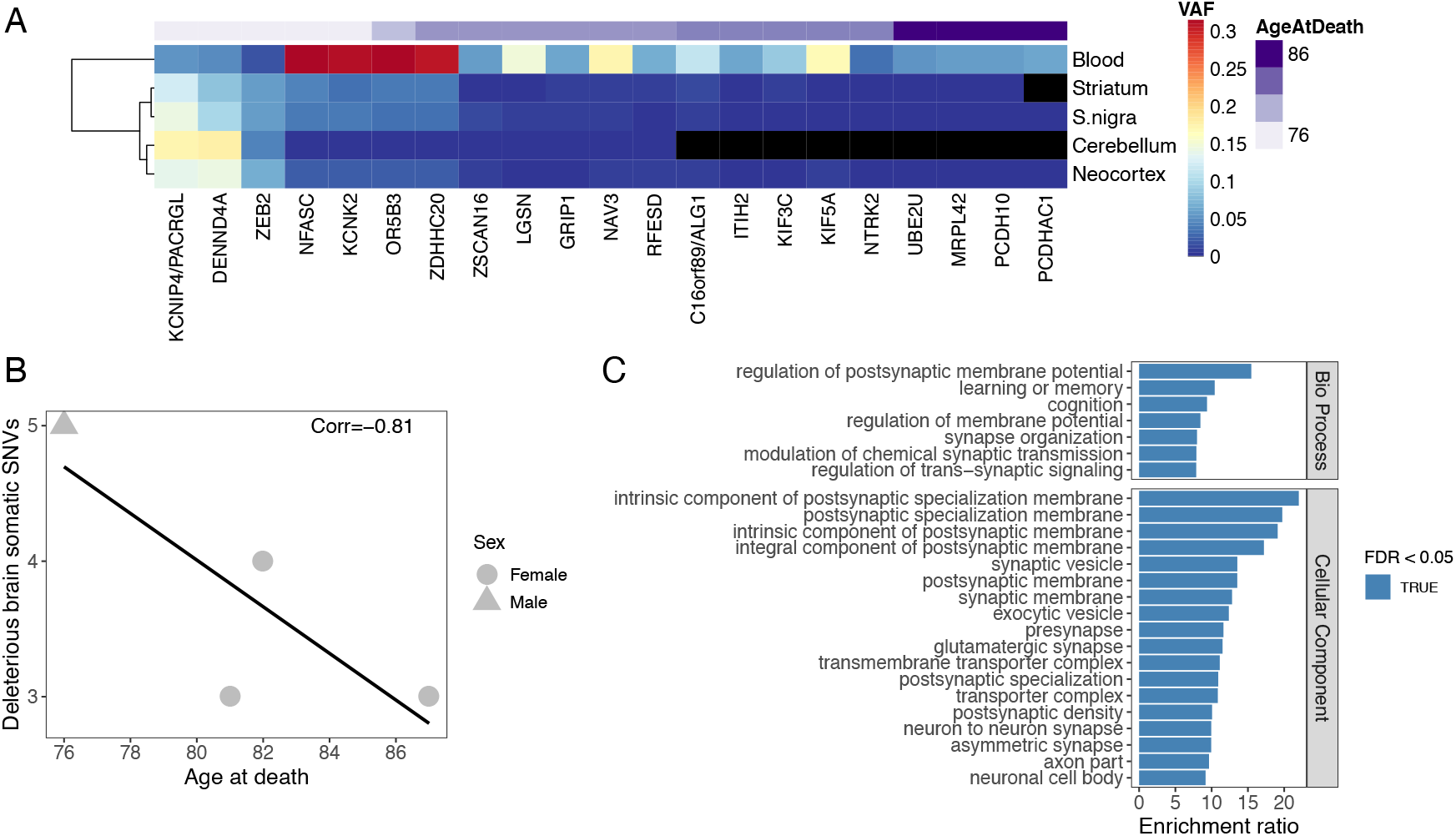
**A. Clustering of tissues by VAF**. Frequency of the 21 somatic variants found in brain was used to cluster the tissues. Genes are shown and variants are ordered by individual, with age at death shown on top. Black tiles indicate the variant did not pass all validation criteria in amplicon sequencing, but could still have support in the tissue. **B. Age correlation**. Correlation between each patient’s number of potentially deleterious variants (nonsynonymous, in splicing consensus sites or in UTRs) found in brain and age at death. The line shows a non-significant fitted linear model (p-value = 0.19). **C. Functional enrichments of extended gene set**. Enrichment ratio of the top 25 terms by FDR of an overrepresentation enrichment analysis of the coexpression network extended gene set. The considered databases were geneontology Biological Process, Cellular Component, Molecular Function; Human Phenotype Ontology and disease GLAD4U.

### Patients with more deleterious sSNVs in brain tend to die earlier

We then tested the relationship between the number of potentially deleterious brain variants and individuals’ age at death. We excluded synonymous sSNVs (4 variants, including the only sSNV validated in DV8’s brain, Supplementary table 4) and intronic variants not affecting donor or acceptor sites (2 variants), leaving 11 nonsynonymous sSNVs, 3 variants affecting splicing consensus sites and 1 variant in a UTR region, as these are more probable to affect age at death. The age of onset was unknown for one of the four individuals included in this analysis, DV1. As age of onset is highly correlated with age at death (Pearson’s r = 0.9), we used it to evaluate the correlation between age and the number of sSNVs. Indeed, counts of deleterious somatic variants identified in brain were negatively correlated with age at death (Figure 4B, Pearson’s r = −0.81) but unsurprisingly from this small data set, the trend was not significant (P-value = 0.19).

### Genes carrying sSNVs in Parkinson are enriched in synaptic processes functions

Out of the 21 sSNVs found in brain, 4 nonsynonymous variants were predicted to be deleterious by SIFT. Remarkably, three of the genes carrying these variants are related to processes relevant in the brain. *GRIP1* is involved in transmission across chemical synapses and the regulation of neuron projection arborization (Geiger et al. 2014). *KCNK2* (or TREK-1) encodes a voltage-independent potassium channel essential in securing saltatory conduction at high frequency on myelinated afferent nerves (Kanda et al. 2019). *UBE2U*, a ubiquitination enzyme, is a candidate regulator of chromatin responses at double strand breaks (Guo et al. 2017) which are of fundamental relevance for gene expression in the brain (Madabhushi et al. 2015). Furthermore, ubiquitination dysfunction has been linked to Parkinson (Geisler et al. 2014) and Alzheimer (Gómez-Ramos et al. 2015). The other variant is in *DENND4A*, a secondary guanine nucleotide exchange factor that activates Rab-10, participating in the insulin-regulated glucose transporter GLUT4 translocation to cell membranes (Sano et al. 2011).

We explored the biological pathways potentially affected by the identified somatic mutations with an ORA. For this analysis we only included the 15 sSNVs that were nonsynonymous changes, that could affect splicing consensus sites or in UTR regions and were validated in a brain tissue. Top enriched terms included “glial cell projection”, “axonogenesis” and similar processes (Supplementary figure 9). However, as expected from such a small set of genes, none of the enrichments were significant (FDR ≤ 0.05). To gain power and retrieve the GO terms that better describe the functional context of the validated sSNVs, we expanded the gene set by adding genes with significantly high coexpression scores (above 900) as reported in the STRING database (Szklarczyk et al. 2019), resulting in a total of 177 genes. Performing an enrichment analysis with the expanded gene set, we found significant results including many synaptic terms (top 25 in Figure 4C), showing that our validated sSNVs affect genes that are tightly connected to the protein networks associated to these functions. Besides the Gene Ontology databases, to test a more direct relationship of our expanded gene list with phenotypes and diseases, we also included the Human Phenotype Ontology and the GALD4U disease databases. Remarkably, chorea, dyskinesia and tremor were among the significantly enriched phenotypes and neurodegenerative diseases and Parkinson disease among the GLAD4U database enriched terms (Supplementary Table 5). To further investigate the tissue specificity of the genes harboring the 15 validated sSNVs, we performed a tissue enrichment using TissueEnrich (Jain and Geetu 2019). In accordance with the functional enrichment, cerebral cortex was the only significantly enriched tissue (adjusted P-value = 0.003 and Supplementary figure 10).

To further explore the consequences of the validated variants, we examined their effect on protein structures and found two interesting cases. All tissues from patient DV7 carried a mutation (p.Gly998Glu) in *KIF5A* (Uniprot Q12840), a kinesin heavy chain protein. Using Genome3D (Lewis et al. 2015), we found a DomSerf (Buchan et al. 2013) structural prediction for the region containing this amino acid (913-1032, confidence 100). The affected residue is in the surface of this small globular C-terminal domain (Supplementary figure 11, left). This region interacts with kinesin adaptor proteins such as TRAK1 and TRAK2, which mediate cargo binding (Hirokawa and Noda 2008), suggesting the possible relevance of this sSNV. All DV1’s tissues but cerebellum carried a substitution in *UBE2U* (Q5VVX9), p.Pro96Ala, which is included in a DomSerf structure of residues 2-157 (confidence 100) and appears to be close to the active site. Although it seems improbable that the structure of the active site changes as a consequence of this mutation (Supplementary figure 11, right), its flexibility or orientation may be affected.

## Discussion

The role of somatic mutations in a variety of diseases is becoming increasingly clear (Gleeson et al. 2000; Messiaen et al. 2011; Poduri et al. 2012; Priest et al. 2016; Bar et al. 2017; Nicolas et al. 2018; Mensa-Vilaró et al. 2019), including neurodegenerative (Park et al. 2018) and neurodevelopmental disorders (Dou et al. 2017). Here, we explored the presence of somatic mutations in a cohort of 10 Parkinson disease patients and validated 27 somatic SNVs whose functions are enriched in synaptic processes, suggesting a potential role of somatic variants in Parkinson disease.

sSNV calling from sequencing data outside the framework of paired samples still poses a challenge. Here we present COSMOS, an approach to accurately identify somatic mutations, that is highly customizable to each experiment’s requirements, COSMOS (https://github.com/ilobon/COSMOS). We highlight the relevance of a panel of individuals to help identify germline variants and recurrent artefacts, which at the sensitivity a reasonable coverage provides, are far more common than somatic mutations. We also demonstrated that having multiple samples from the same individual can help increase accuracy and sensitivity limits, as previously suggested (Kim et al. 2019). Although this advantage can be offset by a panel of individuals for germline variant and recurrent artefacts identification, relevant mutations can be depleted in the context of neurodegeneration, so using unaffected tissues might be key for their discovery. Indeed, some of the somatic variants we validated did not have enough support in the brain tissues, and it was only the information from blood that allowed their detection. This could result from their random amplification in tissue maintenance of blood, especially in older aged individuals who show higher levels of clonal hematopoiesis (Watson et al. 2020) but could also be a consequence of depletion in the central nervous system. Including a wider variety of tissues in future studies could clarify this scenario.

A possible confounding factor in this type of analysis is blood contamination, which would result in the presence of the same somatic mutations at lower frequencies in other tissues. Our results show that 3 of the validated variants have consistently higher VAFs in the brain tissues than in blood. This demonstrates that blood cells are not a subset of the brain bulk samples. It would be interesting to include other tissues in this type of analyses, such as epithelia, which shares the advantage of the random amplification and absence of the putative depletion without this risk of contamination. Nonetheless there is positive selection of sSNVs in normal skin (Martincorena et al. 2015) and the extent to which this can be detrimental for their use as an outgroup tissue remains unknown.

The main caveat of our study is the lack of control individuals, for which paired tissue samples of similar quality are difficult to obtain. However, we took advantage of the remaining 45 samples and used them as a background panel to remove false positives with high statistical power. Also, all validated sSNVs were present in blood, discarding that this is an effect of using brain samples. Finally, their mutational spectrum agrees with our expectations. Extending this experiment to more tissues and individuals will be key to discover the fine role of somatic mutations in Parkinson disease. In addition to somatic mutations, we found that our cohort carried germline variants linked to the disease, hinting at the possibility that the presence of somatic mutations might contribute as an additive factor that increases an individual’s susceptibility or disease severity.

We discarded variants when multiple individuals had reads supporting the same allele. This is because the probability of recurrent errors is much higher than the somatic mutation rate, especially given the number of divisions we can disentangle with bulk medium-coverage sequencing data. However, in the same way germline variants are recurrent because they appear in more fragile or tolerated regions of the genome, somatic mutations will surely be subject to the same processes. Furthermore, variants might be selected for or against in different tissues, as shown for epithelia (Martincorena et al. 2015, 2018; Lee-Six et al. 2019; Yoshida et al. 2020; Moore et al. 2020). Using more sensitive sequencing techniques, such as duplex sequencing (Schmitt et al. 2012), will be fundamental to discover such somatic recurrent events. Finally, exome sequencing data, although more direct in its variant interpretation, is limited to a narrow set of regions of the genome. This limited our power to observe different cell lineages and their presence at each tissue or perform mutational signature deconvolution. Furthermore, the decreasing cost of sequencing and recent advances in contact maps (Lu et al. 2020) make finding and interpreting non-coding variants more accessible.

The somatic mutations we found in Parkinson patients seem to contribute to the disease, as they affect genes involved in neuronal and axonal pathways and interact with genes associated with the disease. Together with previous studies, our results suggest an exciting new research path for the study of diseases, especially complex diseases such as neurodegenerative disorders. This new evidence supports that not only germline point mutations, copy number variants, mitochondrial variants and the environment are relevant, but also somatic mutations can contribute to Parkinson disease, probably by affecting the same key pathways. The study of somatic mutations in larger cohorts can help to identify the relevant molecular routes, helping us understand the disease and finding potential therapy targets.

## Methods

### Exome sequencing

Tissue samples from cerebellum, neocortex, striatum and substantia nigra were collected at patients’ autopsies, 6 to 18 hours after death, and kept frozen at −80 °C at the HCB-IDIBAPS biobank. Blood samples from the same individuals were obtained from stored vials. DNA extractions were carried out with the Qiagen DNeasy Blood & Tissue Kit. Genomic DNA samples were randomly fragmented into 150-200 bp length sequences. Adapters were ligated and the resulting templates were purified with AgencourtAMPure SPRI beads. Libraries were amplified by ligation-mediated polymerase chain reaction (LM-PCR). The Exon Focus SureSelect kit from Agilent was used to capture the exome and paired-end 100 bp sequencing was performed on an Illumina Hiseq2000 platform.

### Bam processing

The resulting FASTQ files were mapped with BWA v0.7.8 mem (Heng Li 2013) to the human hs37d5 assembly. Lane-specific read groups were added with Picard Tools v1.95 (Broad Institute 2013). AddOrReplaceReadGroups and bams were merged by sample with samtools v1.9 (H. Li et al. 2009). Read duplicates were removed with Picard Tools v1.95 MarkDuplicates REMOVE_DUPLICATES=true. Base quality score recalibration and indel realignment were applied following GATK’s best practices (DePristo et al. 2011) with GATK v3.6 (McKenna et al. 2010). Secondary alignments were also excluded with samtools view -F 256.

### Germline variants

Germline variants were called with GATK v3.6. First, GVCFs were obtained for each sample independently with -T HaplotypeCaller -emitRefConfidence GVCF. Then, all samples were genotyped together with -T GenotypeGVCFs and a standard hard filter was applied with -T VariantFiltration --filterExpression “QD < 2.0 || FS > 60.0 || MQ < 40.0 || MQRankSum < -12.5 || ReadPosRankSum < −8.0”. A PCA of the hard-filtered genotypes was performed with EIGENSOFT v7.2.1 (Price et al. 2006). Information on the called variants was annotated with SnpEff 4.3t and SnpSift 4.3t (Cingolani et al. 2012) and dbNSFP was used to add population frequencies, effect prediction and conservation scores. Overrepresentation enrichment analysis was performed with WebGestalt (Zhang, Kirov, and Snoddy 2005) using the Gene Ontology (GO) database (Carbon et al. 2009) for molecular functions and genome protein-coding genes as background.

### Somatic variant calling

Somatic variants were called using HaplotypeCaller and VarScan 2 with lax parameters. Then, an extensive filtering strategy was applied to recover high confidence somatic SNVs. For HaplotypeCaller somatic variant calling, GVCFs per sample were obtained with GATK -T HaplotypeCaller -ploidy 10 -A StrandAlleleCountsBySample -- emitRefConfidence GVCF. Then, all GVCFs were genotyped together per chromosome with -T GenotypeGVCFs -L chr -ploidy 10 -A StrandAlleleCountsBySample to obtain somatic SNV and indel calls. For VarScan 2 (Koboldt et al. 2012) somatic variant calling, mpileup files were obtained with samtools mpileup per individual. Then, single nucleotide variants were called with VarScan v2.3.2 mpileup2snp with lax parameters: --min-coverage 1 --min-reads2 1 --p-value 1 --min-var-freq 0.000001 --output-vcf. Indels were called with mpileup2indel with the same parameters.

Depth of coverage files were obtained with GATK v3.6 DepthOfCoverage and GC content per target was calculated with GCContentByInterval. Then, CNVs were called jointly for all samples with XHMM v1.0 (Fromer et al. 2012) with standard parameters following its recommended best practices. Short tandem repeats in hs37d5 were determined with Tandem Repeats Finder v.4.09 (Benson 1999) with parameters 2 7 7 80 10 12 500 -h. Homopolymers were then extracted based on their homogeneous repeat motif. The 1000GP strict mask FASTA files were obtained from the 1000GP FTP (ftp://ftp.1000genomes.ebi.ac.uk/vol1/ftp/release/20130502/supporting/accessible_genome_masks/) and were transformed into a BED file. BED files for WGAC segmental duplications, common dbSNP SNPs and mappability for 100mers for hg19 – which shares coordinates with hs37d5 – were obtained from the UCSC table browser (Karolchik et al. 2004). This information was annotated in the VCF files with BCFtools (H. Li et al. 2009). A panel of normals (PON) containing allele counts at each genomic position along 428 individuals sequenced at the Sanger Institute was used to identify recurrent errors.

### Somatic variant filtering using COSMOS

We first defined a non-callable set of positions, including off-target calls as well as those overlapping with the 1000GP strict mask, the WCAG track, mappability lower than 1, by homopolymers or within 5 base pairs of indels. Additional sources of artefacts were identified in the lax call sets by manual inspection of the raw data with IGV (Robinson et al. 2011), oftentimes evidenced by recurrent patterns in different individuals. To filter calls we wrote a python module, COSMOS (Combined Or Single sample MOSaicism detection), which is available in https://github.com/ilobon/COSMOS. The first step is to annotate all the information necessary to classify true and false calls. Then, variants are filtered according to the user indicated features and thresholds. Two different filtering approaches can be used. When multiple samples – tissues or replicates – from the same individual are available, their combined information can be used to filter calls more accurately. This is, since true and false variants have partially overlapping values in the determining features, even true somatic mutations can fail some tests. Hence, we can take advantage of the multiple samples, and assuming the variants are present in more than one of them, require than at least *n* samples pass each filter, allowing a different combination of passing samples at each feature (--combined TRUE -- nSamplesPerInd n). COSMOS can also be used to filter each sample individually (--combined FALSE), or output the result of both approaches (--combined BOTH --nSamplesPerInd *n*).

To filter our Parkinson exome data, we used both approaches, requiring at each position either a single sample passing all filters or any combination of four out of the five tissues passing at every filter. The parameters we used were *-c BOTH -ns 4 -ad 2 -adss 3 -vaf 0.5 -dp1 20 -dp2 100 -sr 2 -pr* 4 *-sp 0.05 -b 0.05 -nrl 4 -hap 4 -cnv NO -pir 4,4 -vafq 0.4 -clip 0.9 -mq 0.05 -mm 0.05 -pon 0.05*. In short, this requires at least 2 reads supporting the alternative allele, or 3 for the single sample approach; a variant allele frequency lower than 0.5; a depth between 20 and 100; a strand count ratio <2; a pair count ratio <4; non-significant p-values for a Poisson test of strand counts; a significant binomial test of allele counts; at most 4 variants within read length; less than 4 haplotypes; absence of XHMM CNV call; PIR score of 4, meaning that position in reads of each allele is not biased; a variant allele frequency from direct bam read counts <0.4; an allele clipping ratio difference <0.9; non-significant Mann-Whitney tests for mapping quality an number of mismatches per allele and a significant beta-binomial test for allele counts compared to the PON. More features and different thresholds can be used for filtering depending on each dataset characteristics such as depth.

### Amplicon-based deep sequencing

To validate the high confidence variants, we performed amplicon-based deep sequencing (ADS) by using rhAmpSeq technology (IDT, Coralville, USA). This is a multiplexed strategy that amplifies all selected positions in a unified reaction. We then sequenced the amplified material from each sample in a MiSeq v3 run obtaining paired end 300bp reads to a mean coverage ~18,000X. Data were processed the same way as the exome files. Sequencing data at each position in the samples from other individuals were used as negative controls.

## Data Availability

Sequencing data are available on request and will be deposited in a public repository. Methods including COSMOS are available in GitHub

https://github.com/ilobon/COSMOS

## Notes

### Competing Interest Statement

The authors have declared no competing interest.

### Funding Statement

MINECO (Ministerio de Economia y Competitividad) grants SAF2016-76340-R to E.S.

### Author Declarations

Samples were acquired after death with informed consent from Parkinson patients at Hospital Clinic Barcelona, Spain. Ethical approval for the study was gained from the hospital's Clinical Research Ethics Committee (Reference HCB/2016/0259) in accordance with LIB 14/2007 and RD 1716/2011.

